# Motor network reorganization associated with rTMS-induced writing improvement in writer’s cramp dystonia

**DOI:** 10.1101/2024.09.30.24314652

**Authors:** Noreen Bukhari-Parlakturk, Patrick J. Mulcahey, Michael W. Lutz, Rabia Ghazi, Ziping Huang, Moritz Dannhauer, Pichet Termsarasab, Burton Scott, Zeynep B. Simsek, Skylar Groves, Mikaela Lipp, Michael Fei, Tiffany K. Tran, Eleanor Wood, Lysianne Beynel, Chris Petty, James T. Voyvodic, Lawrence G. Appelbaum, Hussein R. Al-Khalidi, Simon W. Davis, Andrew M. Michael, Angel V. Peterchev, Nicole Calakos

**Author notes:** **Corresponding author:** Noreen Bukhari-Parlakturk, DUMC Box 2900, Bryan Research Building, 311 Research Drive, Durham, NC 27710.

## Abstract

**Background:** Writer’s cramp (WC) dystonia is an involuntary movement disorder with distributed abnormalities in the brain’s motor network. Prior studies established the potential for repetitive transcranial magnetic stimulation (rTMS) to either premotor cortex (PMC) or primary somatosensory cortex (PSC) to modify symptoms. However, clinical effects have been modest with limited understanding of the neural mechanisms hindering therapeutic advancement of this promising approach.

**Objective:** This study aimed to understand the motor network effects of rTMS in WC that correspond with behavioral efficacy. We hypothesized that behavioral efficacy is associated with modulation of cortical and subcortical regions of the motor network.

**Methods:** In a double-blind, cross-over design, twelve WC participants underwent rTMS in one of three conditions (Sham-TMS, 10 Hz PSC-rTMS, 10 Hz PMC-rTMS) while engaged in a writing task to activate dystonic movements and measure writing fluency. Brain connectivity was evaluated using task-based fMRI after each TMS session.

**Results:** 10 Hz rTMS to PSC, but not PMC, significantly improved writing dysfluency. PSC-TMS also significantly weakened cortico-basal ganglia, cortico-cerebellum, and intra-cerebellum functional connectivity (FC), and strengthened striatal FC relative to Sham. Changes in PSC and SPC BOLD activity were associated with reduced dysfluent writing behavior.

**Conclusions:** 10 Hz rTMS to PSC improved writing dysfluency by redistributing motor network connectivity and strengthening somatosensory-parietal connectivity. A key signature for effective stimulation at PSC and improvement in writing dysfluency may be strengthening of intra-cortical connectivity between primary somatosensory and superior parietal cortices. These findings offer mechanistic hypotheses to advance the therapeutic application of TMS for dystonia.

**Highlights:**

- 10 Hz repetitive TMS to somatosensory cortex reduces writing dysfluency in dystonia
- Increased somatosensory cortex activity correlates with reduced writing dysfluency
- In dystonia + sham-TMS, writing dysfluency correlates with cerebellar connectivity.
- 10 Hz rTMS to somatosensory cortex induces reorganization of the motor network

## 1. Introduction

Writer’s cramp (WC) dystonia is a task-specific focal hand dystonia presenting with involuntary muscle contractions involving the hand and arm muscles during the specific task of writing (simple WC) and other fine motor tasks (complex WC) (1). Patients manifest with abnormal, often repetitive, movements and postures of the hand and arm that can be painful and functionally disabling (2). There are no disease-modifying therapies for dystonia, and current treatments provide symptomatic benefit that is short-lived and variable.

Repetitive transcranial magnetic stimulation (rTMS) is a noninvasive brain stimulation technology that in a meta-analysis of 27 prior studies showed some benefit in reducing dystonia symptoms (3). Across these studies, a predictor of benefit was the stimulation site which varied by the dystonia subtype. In dystonias outside of the upper limb, such as cervical dystonia and blepharospasm, some behavioral benefits were reported after TMS to cerebellum (2/6) and anterior cingulate cortex (3/3) respectively. In upper limb dystonia, behavioral benefit was reported after TMS to motor-premotor cortex (PMC) (9/18 studies) or primary somatosensory cortex (PSC) (1/18 studies) (3–5). Although PSC only showed benefit in one study, it is noteworthy that the reported behavioral benefit was more enduring (two to three weeks) than any of the nine PMC studies (3–5). To best resolve whether one target site is superior, a head-to-head comparison of PMC versus PSC target controlling for the other variables among these studies would be necessary.

In addition to the stimulation site, another predictor of TMS benefit was the stimulation parameters. In upper limb dystonia, behavioral benefit was reported after 1 Hz rTMS (6/18 studies), 0.2 Hz rTMS (1/18 studies) and continuous theta burst (TBS) TMS (2/18 studies) while in cervical dystonia and blepharospasm, only TBS-TMS (2/5 studies) and 0.2 Hz rTMS (3/3 studies) showed benefits respectively (3). Overall, some behavioral benefit was reported after TMS in adult focal dystonias but varied by dystonia subtypes with key factors being stimulation site and stimulation parameters.

To improve the clinical efficacy of TMS in dystonia, more clinical studies are needed to directly compare the previously effective stimulation sites and parameters within subject. In addition, advances in our understanding of the brain mechanism mediating TMS benefit irrespective of brain disorder is critical to enable future rational optimization of this promising non-invasive therapeutic modality. In Parkinson’s disease, for example, 10 Hz rTMS to the motor cortex was shown to increase dopamine release from the basal ganglia (4). This study is relevant to dystonia because it demonstrated that 10 Hz rTMS to the motor cortex could modulate subcortical circuitry that is implicated in dystonia as well. In Wilson’s disease, the same 10 Hz rTMS protocol to the motor cortex improved upper limb dystonia suggesting that 10 Hz rTMS to the motor cortex could improve clinical symptoms of upper limb dystonia (5). Taken together, these two studies motivate further exploration of the potential for this frequency in other dystonias. No study to date has tested the role of 10 Hz rTMS in individuals with adult focal dystonias. Collectively, comparative and mechanistic TMS studies are critically needed to guide further refinements in TMS protocols to achieve clinically meaningful and enduring benefits across multiple dystonia subtypes.

In addition to the stimulation site and parameters, an individual’s brain state can be a critical predictor of TMS benefit -a concept referred to as target engagement (5). For instance, in obsessive-compulsive disorder, TMS studies often use cue-triggered symptom provocation to optimize the brain state for engaging the fear-memory reconsolidation network (6). Along these lines, in WC dystonia, a writing task may be useful to prime the writing motor network of the brain implicated in focal hand dystonia.

Here, we aimed to build on prior TMS studies by directly comparing two stimulation sites that have reported efficacy in WC dystonia. We further aimed to understand the relationships between TMS-induced behavioral changes and brain activity by performing functional MRI. The primary hypothesis was that the stimulation site that will show symptom benefit after TMS will modify key subcortical brain regions known to play a role in the dysfunctional motor network of dystonia. In a double-blind, cross-over study design, a 10 Hz rTMS was applied to both PMC and PSC, and compared to Sham rTMS over the course of three independent sessions, allowing for a reliable within-subject comparison of stimulation site efficacy with several critical parameters being held constant.

## 2. Materials and methods

### 2.1 Study design

The study was a double-blind sham-controlled cross-over design with data collected at Duke University Hospital between September 2018 and September 2022. The study was approved by the Duke Health Institutional Review Board (IRB # 0094131), registered on clinicaltrials.gov (NCT 06422104) and performed in accordance with the Declaration of Helsinki. All participants gave written informed consent prior to any study participation. Inclusion criteria were adults (> 18 years), diagnosed with isolated right-hand dystonia by a Movement Disorder Specialist, more than three months from the last botulinum toxin injection, more than one month from trihexyphenidyl medication, and able to sign an informed consent form. Exclusion criteria were any contraindications to receiving MRI or TMS.

### 2.2 MRI data acquisition and preprocessing

All study participants completed a brain imaging scan pre-TMS and those who consented to TMS also completed it after each TMS visit. All anatomical and functional imaging data was collected on a 3 Tesla GE scanner equipped with an 8-channel head coil. The anatomical MRI scan was acquired using T1-weighted echo-planar sequence with the following parameters: voxel size: 256 x 256 matrix, repetition time (TR) = 7.316 ms, echo time (TE) = 3.036 ms, field of view (FOV) = 25.6 mm, 1 mm slice thickness. During fMRI sequences, participants copied holo-alphabetic sentences on an MRI-compatible writing tablet. The sentence writing was performed in a block design alternated by rest blocks (**Fig. 1, task-fMRI panels**). The CIGAL software presented visual writing instructions and recorded participants’ movements during the fMRI scan (7). *<u>Pre-TMS fMRI</u>*: Functional echo-planar images were acquired using the following parameters: voxel size: 3.5 x 3.5 x 4.0mm^3^, flip angle 90°, TR = 2 s, TE = 30 ms, FOV = 22 mm for 37 interleaved slices in ascending order, writing block: 20 s, rest block: 16 s, total: 12 blocks per fMRI. *<u>Post-TMS fMRI:</u>* Functional echo-planar images were acquired with the following parameters: voxel size: 2.0 x 2.0 x 3.0mm^3^, flip angle 90°, TR = 2.826 s, TE = 25 ms, FOV = 25.6 mm for 46 slices. Writing block: 20 s, rest block: 20 s, total: 5 blocks x 6 runs = 30 blocks per fMRI. fMRI images were preprocessed using fMRIPrep (8) as detailed in the supplementary methods. FMRIs with excessive head movements (defined as mean frame-wise displacement > 0.5 mm) were excluded from the study.

**Figure 1:**
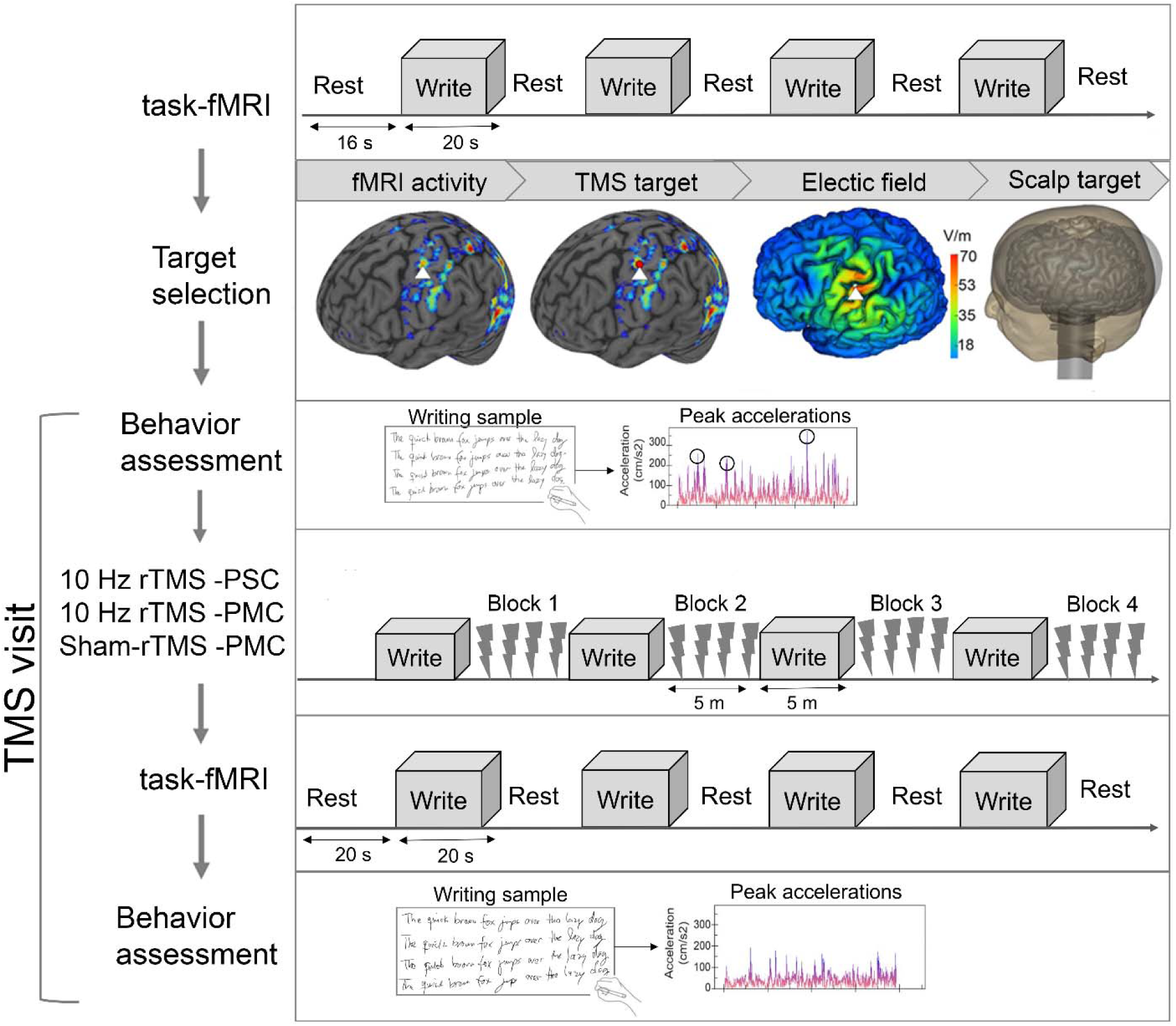
TMS target selection and TMS delivery. To select a scalp target for TMS delivery, each subject completed a task-based fMRI at baseline. An individualized scalp target to motor/premotor cortex (PMC) and primary somatosensory cortex (PSC, represented by a red sphere) was then prepared using fMRI and electric field modeling. Subjects received three TMS conditions: 10 Hz rTMS to PSC, 10 Hz rTMS to PMC and Sham rTMS to PMC (total 4,000 pulses). Each TMS condition was delivered over four stimulation blocks during a single visit. To prime the writing motor network during TMS delivery and circumvent concerns that delivering TMS concurrently during a writing task would compromise stimulation accuracy, an interleaved approach of writing task and stimulation blocks was designed. TMS effect was measured using writing behavior and task-based fMRI.

### 2.3 MRI signal analysis

Pre-processed pre-TMS fMRIs from were input into FEAT analysis in FSL software version 6.0 (FMRIB, Oxford, UK) to generate a subject level statistical map. A general linear model in which writing task timing (block design) was convolved with a double-gamma hemodynamic response function to generate the statistical brain map. To account for head motions, fMRIprep reported regressors (CSF, white matter, framewise displacement, and motion outliers) were regressed out from each statistical map. Spatial smoothing with a Gaussian kernel of full-width half-maximum of 5 mm and temporal high pass filter cutoff of 100 seconds was applied during the FEAT analysis. A participant’s statistical brain map representing the brain (BOLD) activity during the writing task relative to rest on fMRI was then generated and used for TMS targeting. A WC group-level statistical map for the writing-based task-fMRI was also computed by importing all WC participants’ statistical brain maps from their baseline task-fMRI into a mixed-effects FLAME1 model.

### 2.4 Personalized TMS target selection

For each TMS study participant, a two-voxel cortical brain mask was generated for TMS targeting to premotor and primary motor cortices (PMC), and for TMS targeting to primary somatosensory cortex (PSC). To constrain the stimulation target to the PMC and PSC regions, each participant’s statistical brain map was overlayed on the WC group statistical brain map, and anatomical masks for precentral (for PMC) and postcentral gyrus (for PSC) from the Harvard-Oxford MNI atlas and the participant’s anatomical scan. Two consecutive voxels in the anatomic region of left PMC and PSC, with peak activation in the WC participant, and the WC group statistical brain maps and within 1 cm from the scalp surface were selected as the fMRI-guided PMC and PSC target for TMS delivery (**Fig. 1, target selection panel, red sphere represents PSC target**). The fMRI guided cortical brain masks were then used to perform prospective electric field (E-field) modeling on each participant’s scalp as detailed in the supplementary methods. The purpose of the modeling was to identify the TMS coil position and orientation on the participant’s scalp that would maximize the directional E-field in the cortical target of interest perpendicular to the closest gyral wall. This optimal coil setup was then used online with the patient’s T1 to localize and visualize the TMS target in the neuronavigation system (Brainsight, Rogue Research, Canada, version 2.4.9). The final personalized TMS targets at left PMC and left PSC for the 12 WC participants are shown overlayed on the MNI brain (**Fig. 2**).

**Figure 2:**
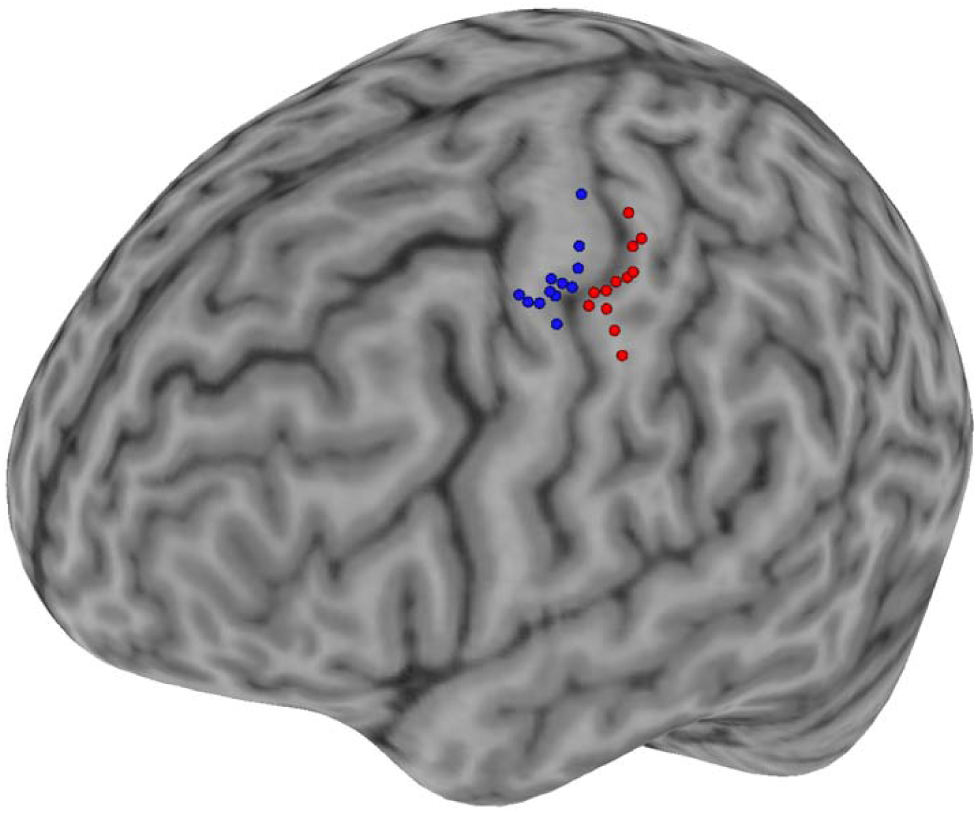
Individualized targets for rTMS to PMC and PSC in WC. The cortical target for rTMS delivery to left PMC (blue) and left PSC (red) was developed using fMRI and electric field modeling. The final target for PMC-TMS and PSC-TMS for each WC participant is shown overlayed on a standard MNI brain.

### 2.5 TMS stimulation

Eligible participants received three TMS visits. The three TMS visits consisted of 10 Hz rTMS to PSC, 10 Hz rTMS to PMC, and Sham rTMS to PMC (**Fig. 1, rTMS panel**). Each rTMS visit was separated by a minimum of one week to allow signal washout. To negate any order effect, each participant was randomized to one of six possible orders for the three TMS conditions. All TMS was performed using an A/P Cool B65 coil attached to a MagVenture R30 device (MagVenture, Farum, Denmark). During each TMS visit, participants first completed an optimal motor cortex localization and motor threshold calculation as detailed in the supplementary methods (9). The active TMS paradigm delivered to each cortical target was 25 trains applied at 10 Hz rTMS with biphasic pulses and an inter-train interval of 10 seconds at 90% resting motor threshold (RMT) for a total of 1000 pulses delivered in a single block (∼five minutes), while participants were in sitting in a recliner. To prime the motor network implicated in focal hand dystonia and circumvent concerns that delivering TMS concurrently during a writing task would compromise stimulation accuracy, an interleaved approach of writing task and brain stimulation was designed. Specifically, each stimulation block was preceded by a writing block (∼five minutes) in which participants performed a sentence copying task. A total of four blocks of TMS alternated with four writing blocks were performed (total 4000 pulses per TMS visit) (**Fig. 1, rTMS panel**). To perform sham stimulation, the same AP coil was used in placebo mode, which produced clicking sounds and somatosensory sensation from scalp electrodes similar to the active mode but without a significant electric field induced in the brain (10). As previously reported, this type of stimulation allows participants to stay blinded during the experiment.

### 2.6 Retrospective TMS coil deviations

During each TMS block, data on the experimental TMS coil location and orientation was recorded every 500 ms in the neuronavigation system and snapped to scalp reconstruction mode prior to exporting it. Data was then imported into SimNIBS software (version 2.0.1 / 3.2.6) to calculate the deviations from the intended TMS coil position and orientation using the retrospective Targeting and Analysis Pipeline (TAP) (11). The TMS coil placement (position and orientation) data were first extruded outwards along the scalp normal by adjusting it for the participant’s recorded hair thickness for each TMS visit. These TMS coil placement data were then used to compute the coil placement deviation from the optimized coil setup constructed during the prospective E-field modeling reported in the supplementary methods. The deviations in the TMS coil placement were calculated in the normal and tangential planes and reported as changes in distance (mm) and angle (degrees). The direct distance between the actual and optimized target was also calculated, as previously reported (11, 12). Due to technical issues with the neuronavigation software, the experimental TMS coil location and orientation during one TMS block was excluded from one participant’s sham-TMS visit.

### 2.7 TMS induced behavior changes

During each TMS visit, participants performed a behavioral writing assay before and after each four-block TMS session (**Fig. 1, behavior assessment panels**). The behavioral assay consisted of participants using a sensor-based pen on a digital tablet (MobileStudio Pro13; Wacom Co, Ltd, Kazo, Japan). Participants copied a holo-alphabetic sentence ten times in a writing software (MovAlyzer, Tempe, AZ, USA). The sensor-based pen recorded the x, y, and z positions and the time function of the participants’ writings. The writing software then transformed the writing samples’ position parameters and time functions using a Fast Fourier transform algorithm to calculate the kinematic features automatically. A previously detailed analysis of these kinematic writing measures showed that the sum of acceleration peaks in a single sentence (henceforth peak accelerations) (**Fig. 1, behavior assessment panel, black circles**) demonstrated high diagnostic potential (sensitivity, specificity, and intra-participant reliability), and associated with patient reported dystonia and disability scales (13). In this study, the peak accelerations measure was used as the primary behavioral outcome measure. Participants performed the behavioral assessment before and after each TMS session. To minimize learning across the three TMS visits, a different holo-alphabetic sentence was used for each sequential visit. The three sentences were: Pack my box with five dozen liquor jugs; The quick brown fox jumps over the lazy dog; Jinxed wizards pluck blue ivy from the big quilt. To measure the change in peak acceleration, each participant’s post-TMS measure was normalized by the mean of their pre-TMS measure using the following equation: [(Post-TMS peak accelerations per sentence)/(mean peak accelerations for all ten sentences pre-TMS)]*100. Higher measures of peak accelerations represent greater writing dysfluency and worsening dystonia. The standard TMS adverse events survey and secondary outcome measures of clinician-rated and participant-reported dystonia scales were also collected before and after each TMS session as detailed in the supplementary methods.

### 2.8 TMS induced fMRI changes

After each TMS visit, participants completed a task-based fMRI. The fMRI task design, acquisition settings, preprocessing and run level analyses are detailed in methods section 2.2 and 2.3 above. To compare changes in BOLD activity across the three TMS conditions, 4D timeseries data were extracted from each fMRI run level FEAT directory using brain masks for regions of interests. The timeseries data for each region of interest was z-scored across runs and analyzed as on-block and off-block writing epochs. To perform functional connectivity analysis, the extracted 4D time series data for each region of interest was correlated pairwise using Pearson’s correlation (R) and Fisher z-transformed.

### 2.9 Brain parcellation and ROI extraction

To compare BOLD activity and functional connectivity analysis across TMS conditions, brain masks corresponding to regions of the motor network previously identified as abnormal in the writing motor circuit of WC dystonia were used in this study. Specifically, an ROI was included if it was reported in at least two of these five isolated sporadic dystonia studies (14–18). Anatomical brain masks were prepared using the Harvard-Oxford MNI brain atlas for the following subcortical regions: left caudate (CAU), left putamen (PUT), left globus pallidus (PAL), left thalamus (THL), left subthalamic nucleus (STN) and left substantia nigra (SN). Since cortex and cerebellum are large brain regions, brain masks for these regions were made using the publicly available Dictionaries of Functional Modes (DiFuMo) brain atlas (19). DiFuMo is a fine grain atlas that parcellated the brain into functional regions of 1024 components, based on data from 15,000 statistical brain maps spanning 27 studies (19). To select DiFuMo brain masks for left superior parietal cortex (SPC), left inferior parietal cortex (IPC), left supplementary motor area (SMA) and right cerebellar lobules VI and VIII (CBL), MNI coordinates from prior neuroimaging studies in dystonia were used (14). For PSC and PMC, each participant’s personalized TMS target (section 2.4) was used as the center to prepare a custom 5 mm spherical mask.

### 2.10 Statistical Analyses

Because the study was a cross-over design with multiple visits and measures, a Mixed-effects Model for Repeated Measures (MEMRM) statistical analysis was used to compare differences in data within participants across the three TMS conditions. For the measure of TMS coil deviations, the MEMRM covariate was TMS condition and since all p-values >0.05, no multiple comparison correction was performed. For the clinician rating scales, the covariates were TMS condition, TMS visit, and clinician rater. For patient rating scales, the covariates were TMS condition, and TMS visit. Since all p-values >0.05, no multiple comparison correction was performed. For the measure of peak accelerations behavior, the covariates for MEMRM were TMS condition, visit, and interaction of TMS condition*visit. Differences across participants for each data set were accounted for by including participant as a random effect variable. Behavior data was adjusted for multiple comparison using Tukey-Holm-Sidak correction (20) with p<0.05 considered significant. For BOLD activity analysis, BOLD activity from 13 brain regions (PMC, PSC, SPC, IPC, SMA, CAU, PUT, PAL, THL, STN, SN, CBL VI, CBL VIII) labeled as motor network were extracted and statistically analyzed using MEMRM. Changes in BOLD activity during the on-block and off-block of writing were analyzed separately. The dependent variable was BOLD activity for each region. The covariates were TMS condition, visit and interaction of TMS condition*visit. BOLD activity analysis was corrected for 60 MEMRM tests using the FDR correction method of Benjamini-Hochberg (21) and p<0.05 considered significant. For functional connectivity, the 13 brain regions were correlated pairwise across the motor network. To focus on clinically meaningful differences induced by TMS, functional connectivities (FC) that showed at least more than minimal effect sizes (defined as Cohen’s D ≥ |0.2|) between active and sham conditions were analyzed using MEMRM test with a setup similar to BOLD activity analysis. FC data were FDR corrected for 126 FC tests for PSC vs. Sham and 166 FC tests for PMC vs. Sham with p <0.05 considered significant. To perform BOLD activity-behavior correlations within participant, BOLD activity from brain regions reported in Table 1 were correlated within participant with their measure of normalized peak accelerations behavior using Pearson’s (R). BOLD activity-behavior correlations (R) greater than or equal to 0.6 for at least one TMS condition were presented. An exploratory posthoc analysis was also performed to compare differences in FC-behavior correlation across the three TMS conditions. FC-behavior relationships in the motor network with R-value greater than or equal to |0.6| for at least one TMS condition were identified and presented in a heatmap (Figure 9B). Using the correlations in Figure 9B, a subset analysis was performed to statistically compare the FC-peak accelerations correlations that differentiated the effective PSC stimulation site from the non-effective PMC and sham conditions. To evaluate for statistical differences, a generalized linear regression analysis (22) was performed for each FC-behavior relationship that differentiated PSC-TMS from the other two conditions. The dependent variable was peak accelerations behavior, the covariates were FC, TMS condition, and interaction term (FC*TMS condition). P-values for the interaction term for PSC-TMS (PSC-TMS condition*FC) across the selected FCs were corrected for multiple comparisons using the Benjamini-Hochberg method(21) with p<0.1 considered significant.

**Table 1:** Changes in functional connectivity induced by PSC-TMS compared to sham-TMS in the motor network.

| Brain Regions | Functional connections | FC mean z-score (SE) |  | Mixed-effects Model for Repeated Measures |  |  |  |
| --- | --- | --- | --- | --- | --- | --- | --- |
|  |  | Sham | PSC | PSC vs. Sham Mean FC difference | SE | t-ratio | p-value* |
| Cortical-striatal | PMC-CAU | 0.49 (0.03) | 0.33 (0.03) | -0.17 | 0.04 | -4.49 | 0.015 |
|  | PSC-CAU | 0.23 (0.03) | 0.10 (0.03) | -0.13 | 0.03 | -4.17 | 0.015 |
| Cortico-cerebellar | SPC-RCBL-VIII | 0.44 (0.03) | 0.27 (0.04) | -0.15 | 0.05 | -3.38 | 0.048 |
| Subcortico-subcortical | PAL-CBL-VI | 0.13 (0.04) | 0.22 (0.04) | +0.17 | 0.05 | 3.61 | 0.041 |
|  | PUT-SN | 0.48 (0.04) | 0.59 (0.03) | +0.16 | 0.04 | 3.53 | 0.042 |
| Intra-cerebellar | R-CBL VI-VIII | 0.51 (0.04) | 0.40 (0.05) | -0.18 | 0.04 | -4.28 | 0.015 |
|  | R-CBL VI-VIII | 0.45 (0.03) | 0.30 (0.05) | -0.20 | 0.05 | -4.12 | 0.015 |

PMC: premotor cortex; CAU: caudate, PSC: primary somatosensory cortex; SPC: superior parietal cortex; R-CBL-VIII: right cerebellum lobule VIII; PAL: pallidum; PUT: putamen; SN: substantia nigra; R-CBL VI: right cerebellum lobule VI; **SE**: Standard Error, \*p-values after MEMRM and FDR correction.

## RESULTS

### Clinical characteristics

Thirty-four participants were assessed for study eligibility (**Fig. 3**). Of those assessed for eligibility, 24 WC participants met the inclusion/exclusion criteria to participate in the TMS study and completed a baseline fMRI before the first stimulation visit. The baseline fMRI from five participants were excluded due to other neurological disorder, structural abnormalities on MRI brain or excess head motion. FMRIs from 19 WC participants were then used to identify a group targeting approach for TMS. Of these 19 participants, 14 consented to participate in the TMS research visits. Two participants who consented to TMS visits were taking a medication that increased the risk of seizures and, therefore, were excluded from undergoing TMS. Twelve WC participants (11/1 males/female; mean age 55 [SD 12.91] years) completed all three TMS visits. Due to technical issues during data collection, one participant’s TMS visit (Sham) was removed from data analysis. Thus, data from 12 WC participants and 35 TMS visits (12 participants x 3 conditions – 1 visit = 35) were used for all analyses in this study. None of the 12 WC participants reported any TMS adverse side effects. The mean symptom duration for the 12 WC participants was 16.4 years [SD 15.54].

**Figure 3:**
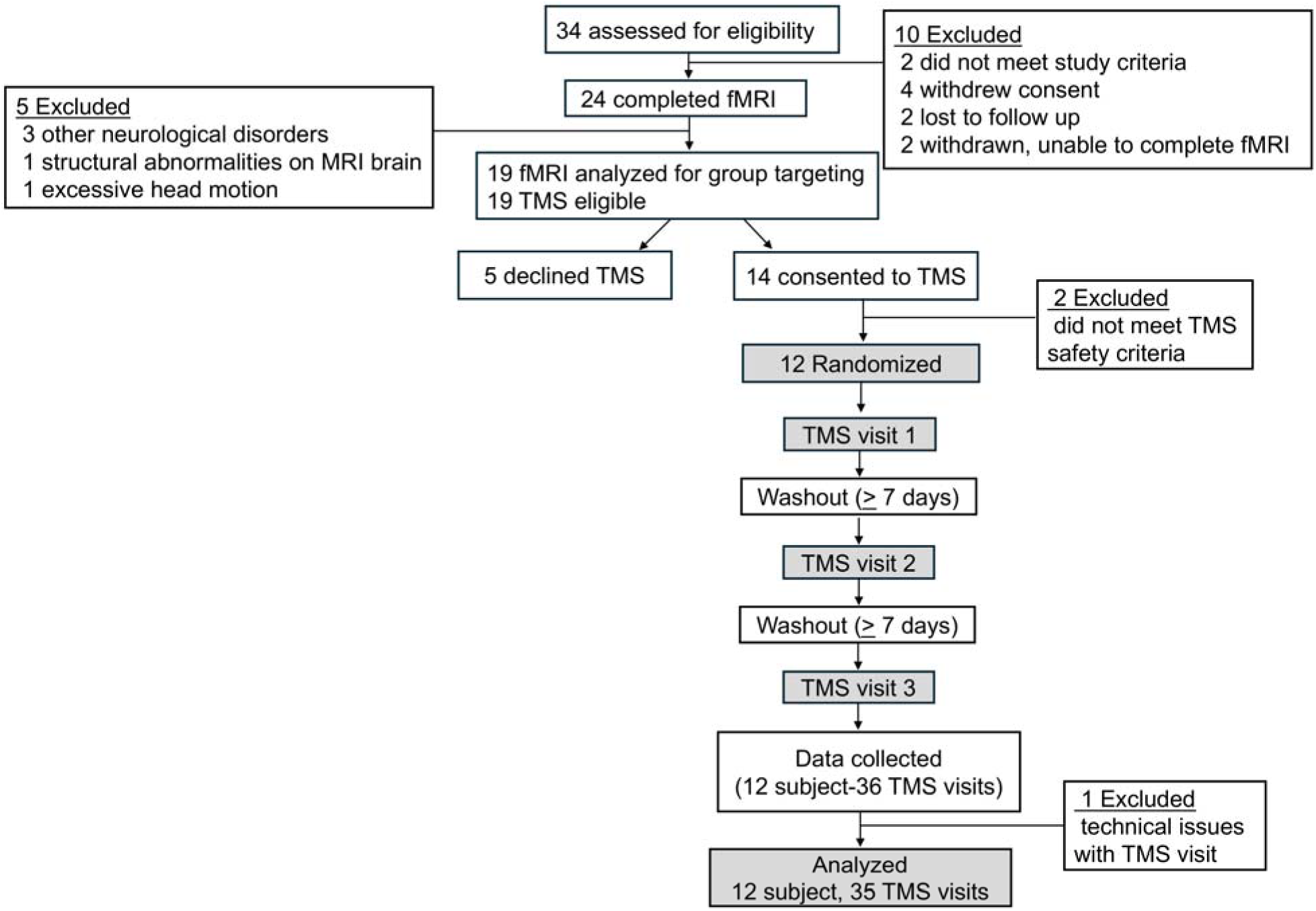
Consort diagram showing the recruitment, inclusion and exclusion number of participants. Total of 34 WC dystonia participants were screened. 24 participants completed fMRI, 14 consented to the TMS study. Of these, 12 participants were randomized and completed the TMS study.

**Figure 4:**
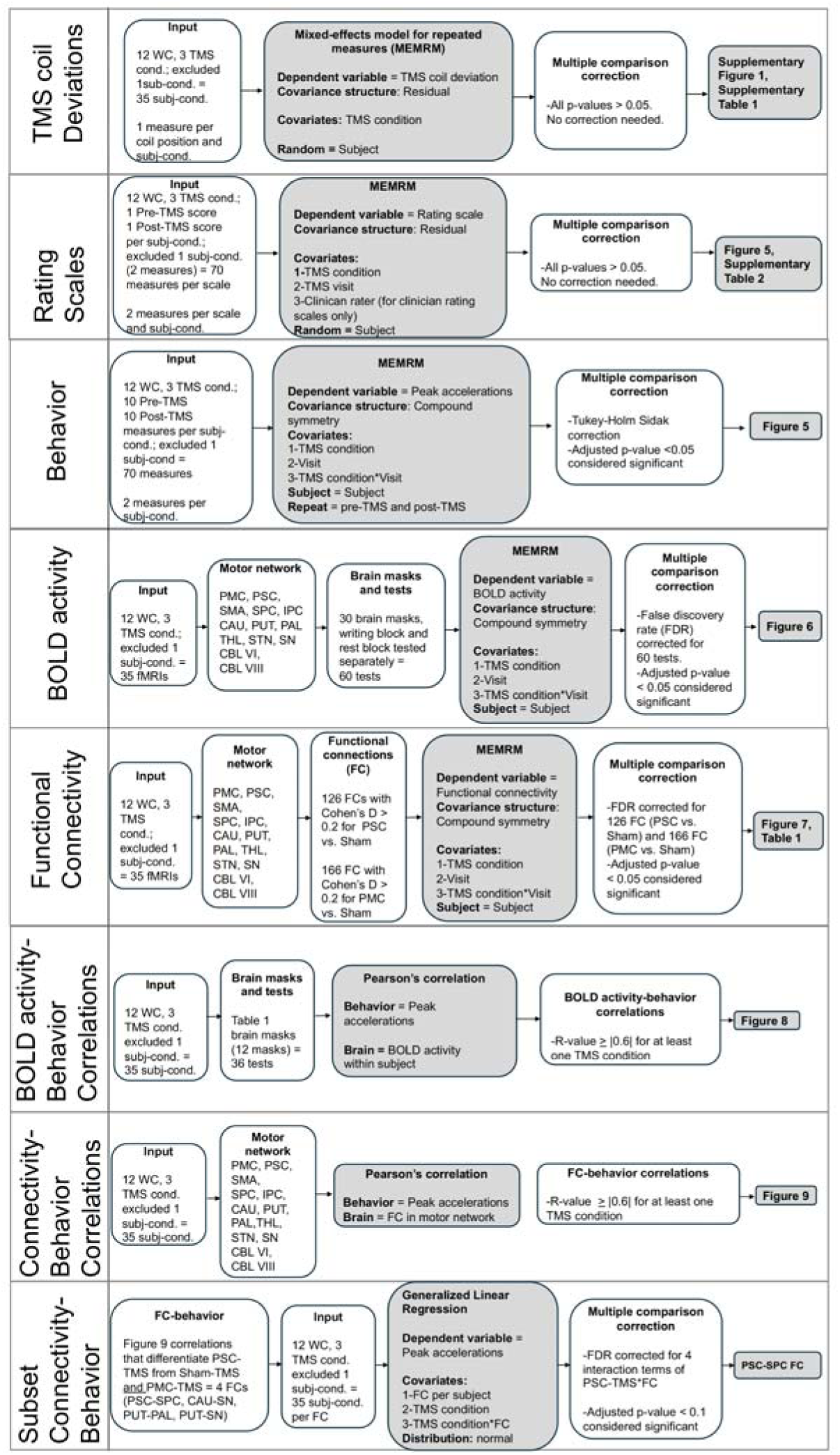
Schematic overview of study data and analytical pipeline. The diagram illustrates the key datasets collected and analyzed in the study. For each dataset, the statistical models, multiple comparison correction methods, and the corresponding figure and/or table resulting from the dataset are reported. Mixed-effects model for repeated measures (MEMRM) was employed with covariates and covariance structures tailored to each dataset. Correlations were performed using Pearson’s R to understand the relationship between peak accelerations, BOLD activity, and functional connectivity. Multiple comparison corrections were performed where applicable.

### No differences in TMS technical delivery

To evaluate the technical delivery of TMS, the position and orientation of the TMS coil during the three conditions were analyzed retrospectively. There were no significant differences in the position or orientation of the TMS coil across the three conditions (**Supplementary Figure 1 and Supplementary Table 1**). Therefore, TMS delivery across the three conditions was technically comparable.

### 10 Hz rTMS to PSC, but not PMC, improved writing dysfluency

In a within-participant comparative design, in which all participants received stimulation to both sites and at the same frequency, we found that 10 Hz rTMS to PSC significantly decreased writing dysfluency compared to Sham-TMS (**Fig. 5A**) [PSC: mean 96.43, SE 1.39; Sham: mean 100.06, SE 0.76; PSC vs. Sham: -1.73, SE: 0.41, t(21): -4.22, p = 0.001] and PMC-TMS [PMC: mean 99.00, SE 0.90; PSC vs. PMC: -1.28, SE 0.40, t(21): 3.23, p = 0.012]. TMS to PMC did not show significant differences in writing dysfluency compared to Sham [PMC vs. Sham: -0.45, SE 0.41, t(21): -1.09, p = 0.639]. These results confirm prior studies that TMS can modify behavior in WC dystonia and show the first results using 10 Hz frequency, within-participant site comparisons, and delivery under a task-primed brain state. Across the clinician rating (BFM right arm dystonia, and WCRS movement score) and participant reported scales (BFM writing score, and ADDS), there were small but consistent improvements in dystonia symptoms after PSC-TMS compared to Sham (**Figure 5B-E, Supplementary Table 2**), “Difference” column, positive values represent improvement, and negative values represent worsening dystonia). However, the effect sizes of these categorical rating scales were small with large variability resulting in no statistical differences across the three TMS conditions.

**Figure 5:**
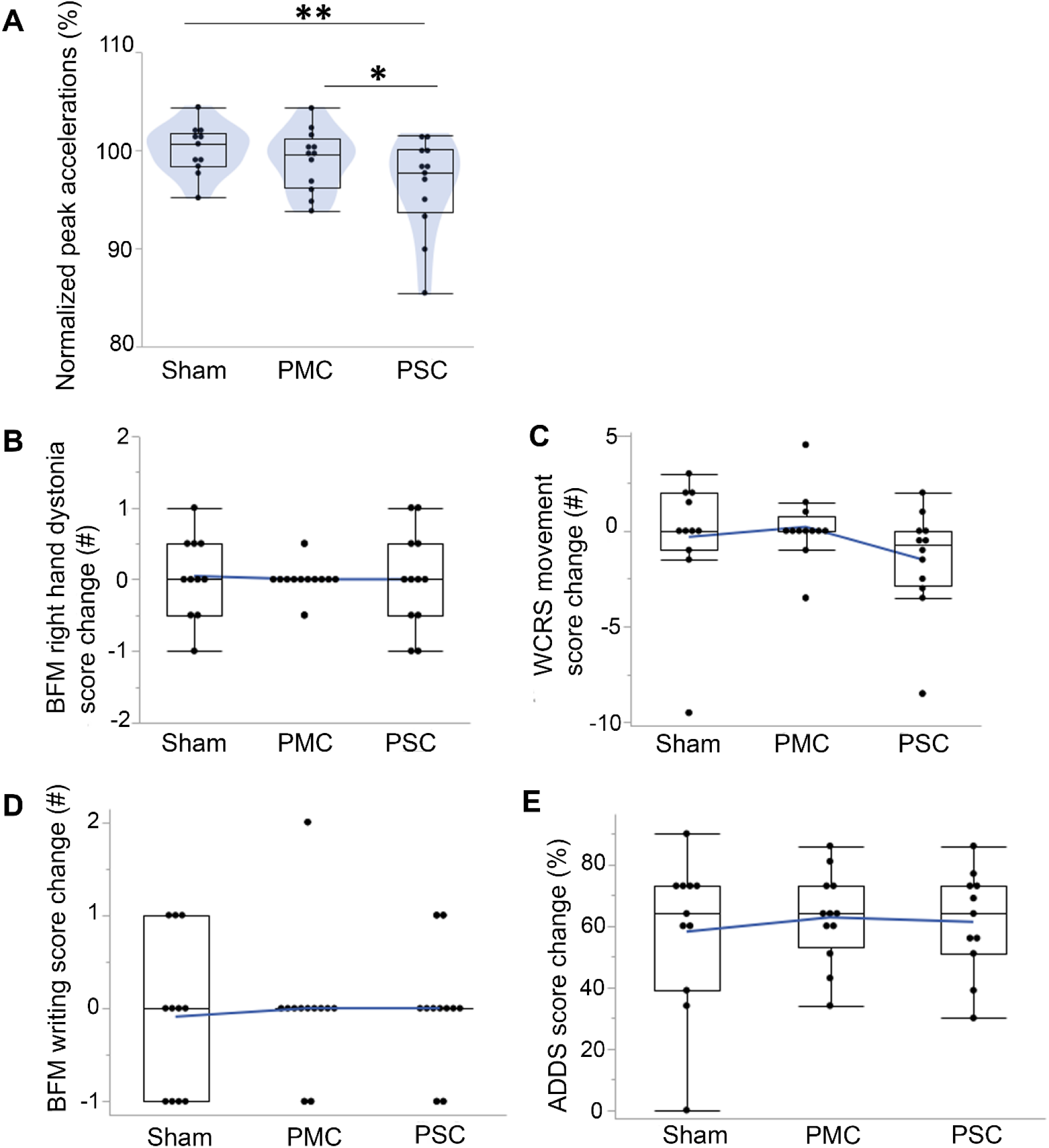
10 Hz rTMS to PSC, but not PMC, reduced writing dysfluency in WC. **A)** 10 Hz rTMS to PSC significantly reduced a measure of writing dysfluency called peak accelerations compared to sham-TMS in a within-subject analysis in WC participants. PMC-TMS did not show significant differences in writing dysfluency compared to sham-TMS. Each data point represents the mean change in peak accelerations for each TMS condition with higher measures representing worsening writing dysfluency. **B-E)** The effect of TMS on right hand dystonia in WC participants was also compared using the clinician-rated scales of B) Burke Fahn Marsden (BFM) right hand dystonia and **C)** Writer’s Cramp Rating Scale (WCRS) movement scores. TMS effect on WC participants’ right-hand disability was reported using the **D)** BFM handwriting disability score and **E)** Arms Dystonia Disability Scale (ADDS). All rating scales were performed before and after each TMS condition. Each data point on the graph represents a subject’s score change (Post-TMS minus Pre-TMS) in the scale. **p<0.01, *p<0.05 after MEMRM test and Tukey-Holm Sidak correction.

### 10 Hz rTMS to either PMC or PSC decreased subcortical activity in the motor network compared to sham-TMS

Considering the differential effect of the stimulation site on behavioral outcomes, we examined how active TMS at these two target sites affected brain BOLD activity during a writing task relative to both rest and sham-TMS condition (**Fig. 6A**). Across the motor network, active stimulation at either of the two TMS target sites showed similar patterns of brain activation during the writing task compared to sham-TMS (**Fig. 6B**). Specifically, 10 Hz rTMS decreased subcortical brain activity and increased BOLD activity at the superior parietal cortex during writing compared to sham-TMS. In the cerebellum, the brain activation pattern during writing varied by the active TMS target site. Active PMC-TMS decreased BOLD activity in lobules VI and VIII while active PSC-TMS decreased BOLD activity in lobule VI only during writing compared to sham-TMS. Overall, these results suggest that cortically delivered 10 Hz rTMS decreased deep brain activity in the motor network compared to sham-TMS with an overall activation pattern during writing that was similar across the two stimulation sites. Changes in BOLD activity after 10 Hz rTMS, therefore, do not fully explain the differential effect of stimulation site on behavioral outcomes.

**Figure 6:**
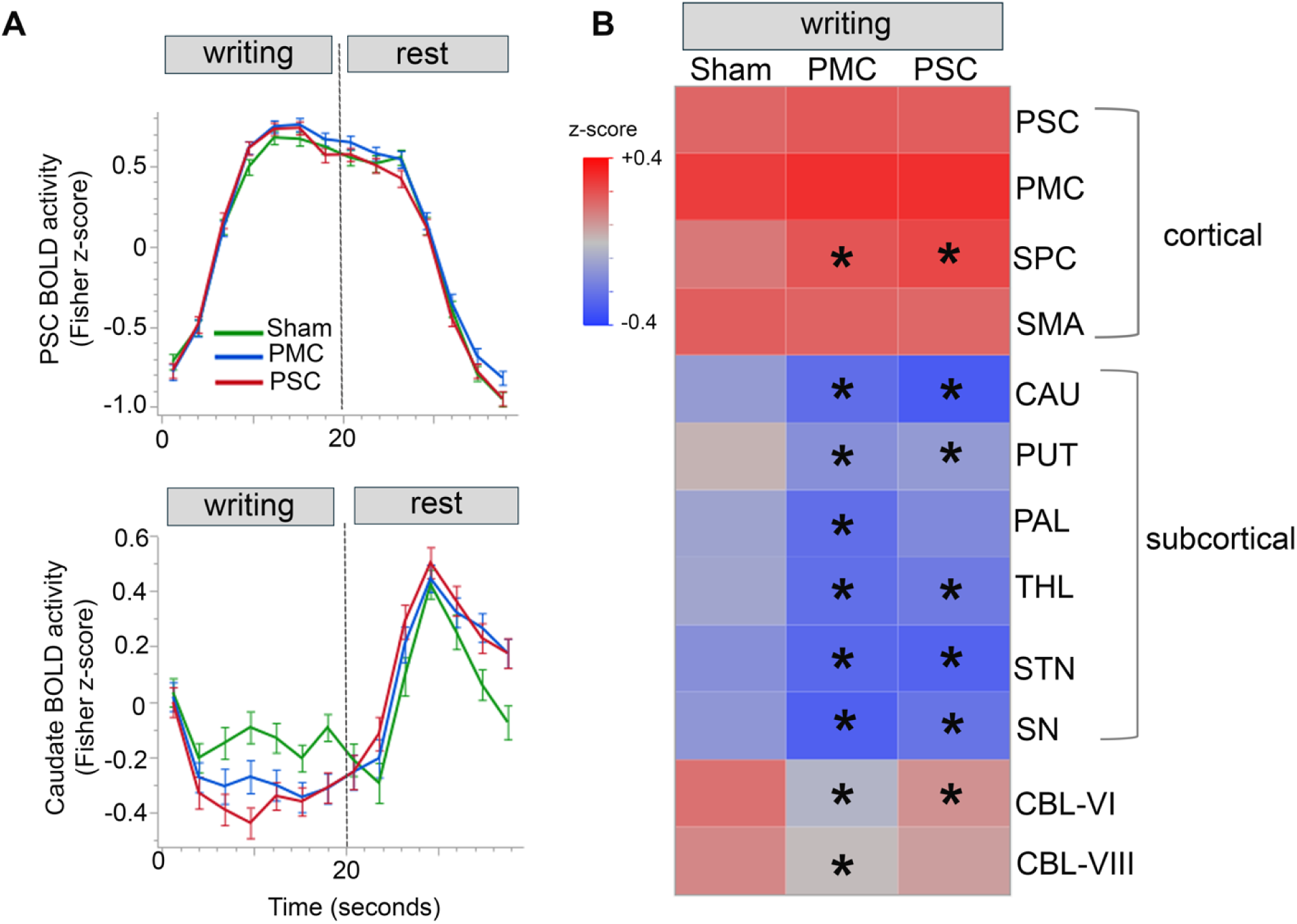
10 Hz rTMS to either PMC or PSC decreased subcortical activity in the motor network. **A)** Graphs represent mean BOLD activity during the writing and rest blocks after each TMS condition [sham-TMS (green), PMC-TMS (blue) and PSC-TMS (red)]. The mean BOLD activity is presented for brain regions of the primary somatosensory cortex (PSC, top graph) and caudate (bottom graph, PSC vs. Sham: -0.19, p <0.0001, PMC vs. Sham -0.14, p < 0.001, PSC vs. PMC: 0.05, p = 0.14, MEMRM and FDR-corrected at p < 0.05). **B)** Heatmap represents mean BOLD activity during the writing block for subregions of the motor network across the three TMS conditions. Asterisks indicate significant difference in mean BOLD activity for PMC-TMS or PSC-TMS compared to sham-TMS (MEMRM and FDR-corrected at p < 0.05). SPC: superior parietal cortex, SMA: supplementary motor area, CAU: caudate, PUT: putamen, PAL: pallidum, THL: thalamus, STN: subthalamic nucleus, SN: substantia nigra, CBL-VI: cerebellum, lobule VI and CBL-VIII: cerebellum, lobule VIII.

### PSC-TMS modified subcortical connectivity, distinct from PMC-TMS and sham-TMS

We next considered if the behavioral outcome differences between the stimulation sites also corresponded to changes in functional connectivity (FC) after TMS. **Figure 7** illustrates the FC changes induced by PSC-TMS compared to sham-TMS. In general, PSC-TMS weakened cortico-striatal FC compared to sham-TMS (thin lines: PMC-CAU, PSC-CAU), cortico-cerebellar FC (SPC-CBLVI), and intra-cerebellar FC (CBL VI-VIII). PSC-TMS also strengthened striato-cerebellar FC (thick lines: PAL-CBL-VI) and striato-nigral FC (PUT-SN) compared to sham-TMS (**Table 1**). There were no FCs that showed significant differences between PMC-TMS compared to sham-TMS. Overall, these findings demonstrate two important points. 10 Hz rTMS to PSC interleaved with writing task predominantly changed subcortical FC compared to sham-TMS. Second, changes in FC induced by TMS may explain the differential effect of stimulation site on behavioral outcomes.

**Figure 7:**
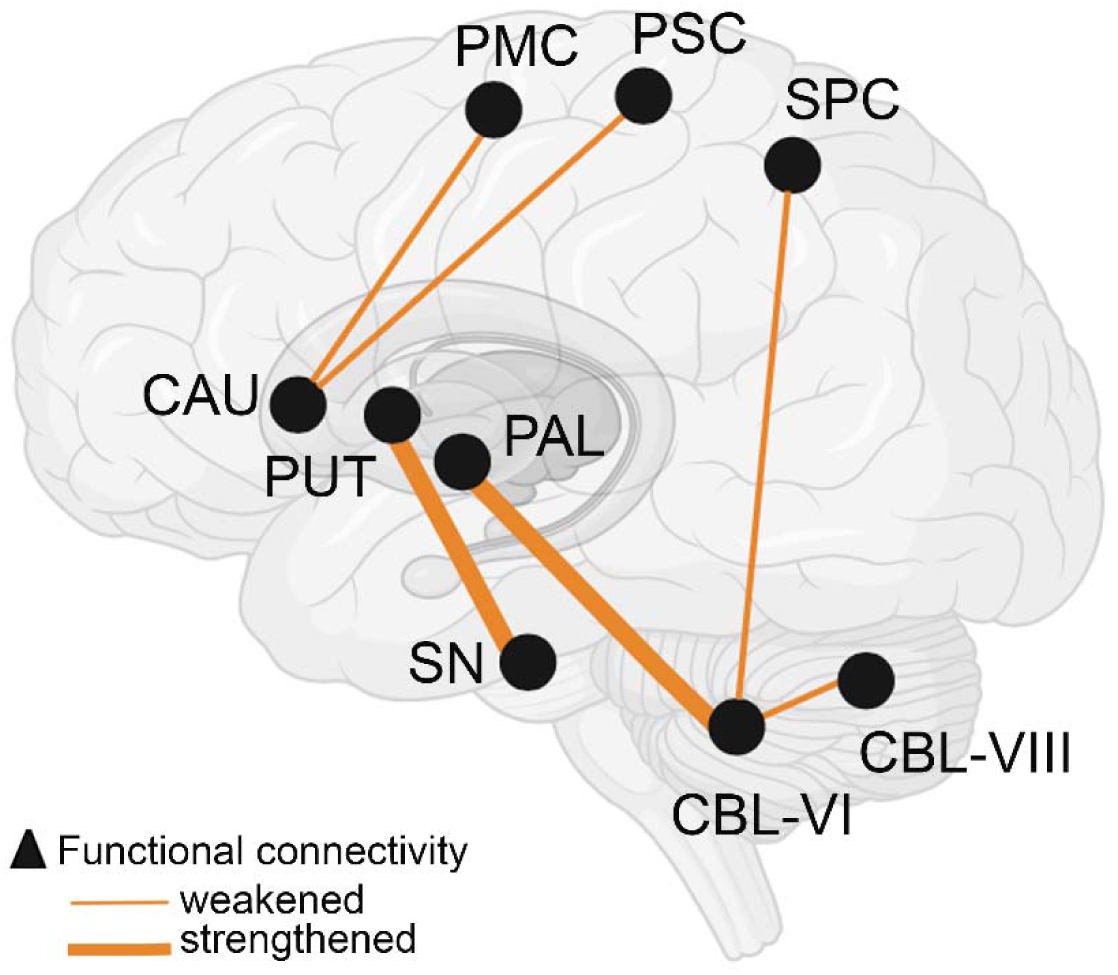
10 Hz rTMS to PSC but not PMC induced changes in cortical-subcortical connectivity in the motor network compared to sham-TMS. The brain model represents TMS induced changes in functional connectivity (FC) in the motor network. Black circles represent brain regions in the motor network and connecting lines indicate FC changes between these regions. Line thickness denotes the direction of FC change (thin line: weakened FC, thick line: strengthened FC). Only FC that were significantly different between PSC-TMS and sham-TMS (p < 0.05, MEMRM with FDR correction) are shown. No significant FC changes were observed after PMC-TMS compared to sham-TMS. Detailed FC values for PSC-TMS versus sham-TMS are provided in Table 1. PMC: premotor cortex; PSC: primary somatosensory cortex; SPC: superior parietal cortex; CAU: caudate; PUT: putamen; PAL: pallidum; SN: substantia nigra; R-CBL VI: right cerebellum lobule VI; R-CBL-VIII: right cerebellum lobule VIII.

### TMS-induced changes in PSC and SPC BOLD activity were associated with reduction in writing dysfluency

We next asked if there was a relationship between TMS-induced brain activation and behavior changes and if this relationship was dependent on the stimulation site. A BOLD activity-behavior correlational analysis was performed for all brain regions in **Table 1** that showed significant differences in functional connectivity between PSC-TMS and sham-TMS. Among these brain regions, PSC and SPC BOLD activity correlated with behavior of peak accelerations. Specifically, increase in PSC BOLD activity after PSC-TMS was associated with reduction of peak accelerations behavior in WC participants (R = -0.84, p = 0.02) (**Fig. 8**). In contrast, there were no correlations observed between PSC BOLD activity and behavior after PMC-TMS (R = -0.39, p = 0.75) or sham-TMS (R = 0.15, p = 0.76). In contrast, increase in SPC BOLD activity correlated with increase in peak accelerations behavior after Sham-TMS in WC participants (R= 0.74, p = 0.01). This SPC BOLD-behavior correlation was not observed after PMC-TMS (R = -0.19, p = 0.55) or PSC-TMS (R = -0.15, p = 0.64). Collectively, these results suggest that TMS induced changes in the association between brain activity and behavioral measures were dependent on the stimulation site and TMS induced activation of PSC and SPC after PSC-TMS were associated with reduction in writing dysfluency.

**Figure 8:**
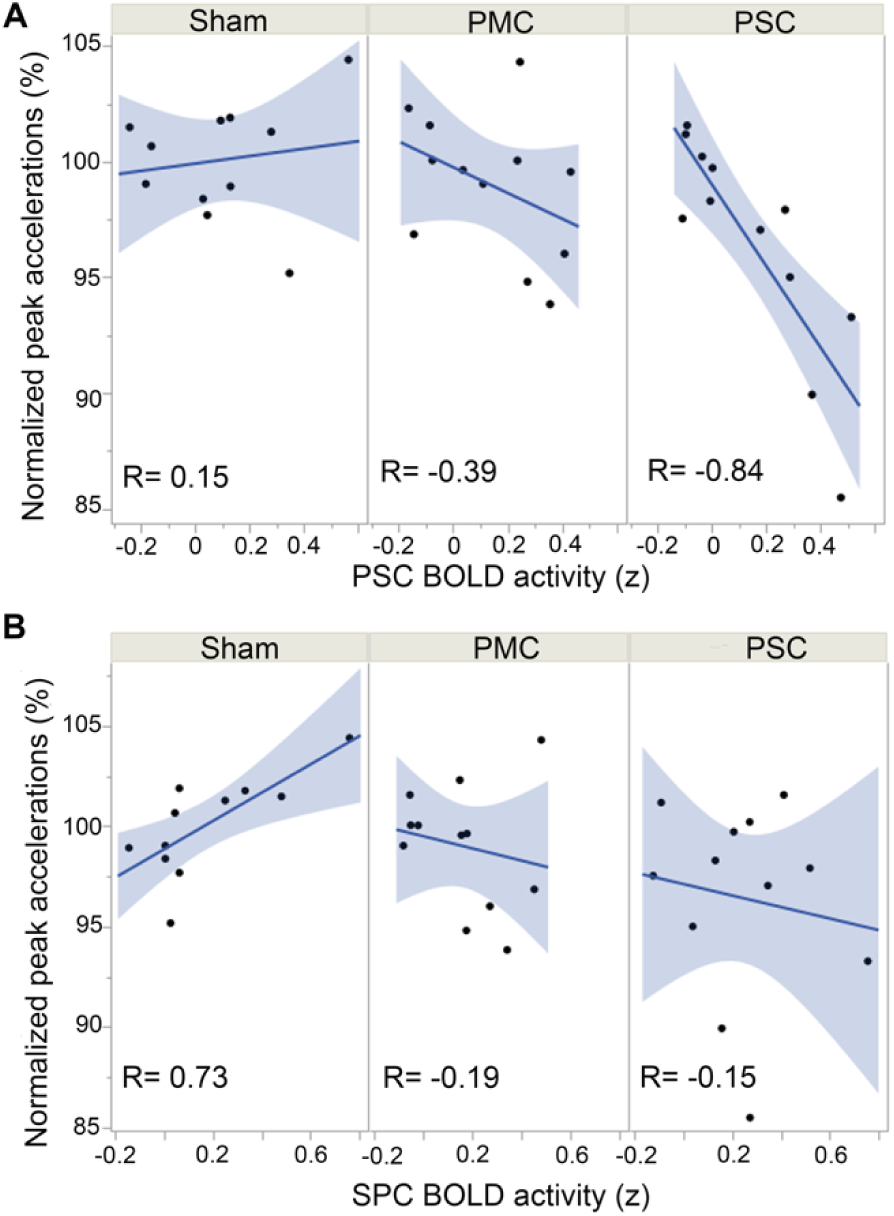
TMS-induced change in BOLD activity at PSC and SPC correlated with reduced writing dysfluency. Graphs represent the correlation between BOLD activity at A) primary somatosensory cortex (x-axis) and B) superior parietal cortex (x-axis) with behavior of peak accelerations (y-axis) for the three TMS conditions. Each data point represents the correlation between a WC participant’s regional BOLD activity and peak accelerations behavior for each TMS condition. Shaded blue regions represent the confidence region for the fitted lines. A Pearson’s correlation (R) is reported for each BOLD activity-behavior correlation and TMS condition.

**Figure 9:**
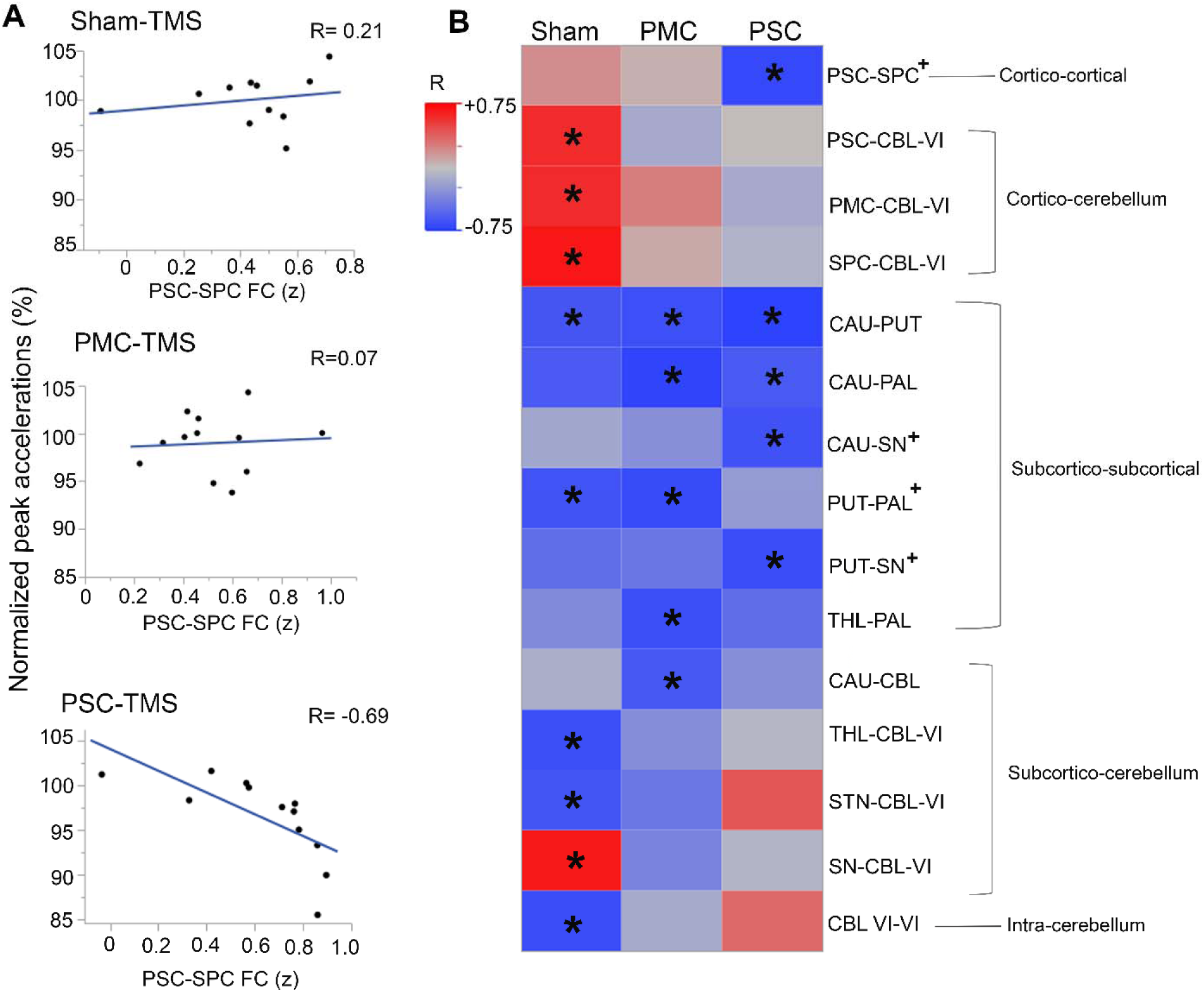
Reorganization of the motor network connectivity after PSC-TMS correlated with reduced writing dysfluency. **A)** Graph represents the correlation between functional connectivity (FC) (x-axis) and peak accelerations behavior (y-axis) for the 12 WC subjects and three TMS conditions. Each data point represents a WC participant’s FC-behavior relationship. The representative scatter plots are organized by TMS condition and shown for the relationship between PSC-SPC FC to peak accelerations behavior. WC subjects showed no FC-behavior correlation for conditions of sham-TMS (R = 0.21) or PMC-TMS (R = 0.07). But there is an inverse correlation between PSC-SPC FC and peak accelerations behavior after PSC-TMS (R = -0.69). **B**) A heatmap of the mean correlation between FCs in the writing motor network and peak accelerations behavior. Each box represents the mean correlation (R) between peak accelerations behavior and a FC for each TMS condition (red box = positive FC-behavior correlation, blue box = negative FC-behavior correlation). Heatmap is reported only for FCs that show R ≥ |0.6| for at least one TMS condition (indicated by an asterisk) and compared with the other two TMS conditions. A subset FC-behavior analysis was performed to compare the four FC-behavior relationships that differentiate PSC-TMS from both sham-TMS and PMC-TMS (indicated by “+”). Across the four FC-behavior relationships, strengthening of PSC-SPC connectivity correlated with significant reduction in peak accelerations behavior after PSC-TMS compared to sham-TMS (PSC vs. Sham: -14.6, p = 0.075, generalized linear regression, and FDR correction).

### PSC-TMS induced reorganization of motor network connectivity was associated with reduced writing dysfluency

Lastly, in a post-hoc analysis, we explored if there was an association between TMS-induced changes in functional connectivity in the motor network and TMS-induced changes in behavior and if this association was dependent on the stimulation site. From this analysis, we made three observations (**Fig. 9**). First, under Sham-TMS condition (control condition), nine significant FC relationships with writing dysfluency were identified with majority of them in connection to the cerebellum (cortico-cerebellum, subcortical-cerebellum, intra-cerebellum). Second, compared to sham, correlations between writing dysfluency and FC involving the cerebellum were no longer present following 10 Hz rTMS to either PMC or PSC. The loss of FC-behavior correlations in these regions was observed to a greater extent after PSC-TMS than PMC-TMS. Third, using the FC-peak accelerations correlations in Fig. 9B, a subset analysis was performed to statistically compare the FC-peak accelerations relationships that differentiated the effective PSC stimulation site from the non-effective PMC and sham conditions. PSC-TMS differed from the other two stimulation sites in four FC-behavior correlations (PSC-SPC, CAU-SN, PUT-PAL, PUT-SN) that spanned cortical and subcortical brain regions (**Fig. 9B, indicated by “+” sign**). Of these four correlations, PSC-TMS significantly differed from sham-TMS in the PSC-SPC FC-behavior association (PSC vs. Sham: -14.6, p = 0.075, generalized linear regression, Benjamini-Hochberg adjustment for multiplicity) (**Fig. 9A**). Collectively, these results demonstrated that reduced writing dysfluency after PSC-TMS may be mediated by loss of functional connectivity-behavior associations to the cerebellum, and/or gain of functional connectivity-behavior associations to cortical and subcortical brain regions. Furthermore, the association between writing dysfluency and intracortical connectivity (PSC-SPC) may be a key signature for PSC-TMS induced brain-behavior changes.

Overall, TMS target comparison demonstrated that 10 Hz rTMS to primary somatosensory cortex but not premotor cortex significantly changed functional connectivity and markedly redistributed functional connectivity-behavior associations that spanned cortical and subcortical regions of the motor network.

## DISCUSSION

The present study compared the efficacy of two TMS cortical sites in reducing dystonic behavior, each previously shown to be beneficial in separate dystonia studies, and aimed to identify a TMS-induced brain mechanism underlying the observed behavioral improvement. We report three key findings. First, 10 Hz rTMS to primary somatosensory cortex significantly reduced writing dysfluency compared to Sham and to 10 Hz rTMS at the premotor cortex. These results suggest the clinical potential of 10 Hz rTMS to the PSC for this rare brain disorder. Second, we identified that TMS to the same region may have improved behavior by changing subcortical connectivity in the motor network. Third, we demonstrate that the intra-cortical connectivity between primary somatosensory and superior parietal cortices are a key predictor for effective stimulation at PSC. Collectively, the present study findings will guide future refinements in TMS protocols to achieve clinically meaningful and enduring benefits in this rare brain disorder.

Our first principal study finding was that 10 Hz rTMS to PSC significantly reduced writing dysfluency in WC dystonia. The twenty-minute PSC-TMS session showed an effect size of 0.96 compared to Sham. This effect size is among the highest reported for TMS studies in dystonia. The kinematic writing metric was selected based on its high diagnostic performance in a prior exploratory study comparing 22 kinematic writing measures from writer’s cramp and healthy volunteers (13). In that study, across the 22 kinematic measures, peak accelerations showed high sensitivity, specificity, intra-subject reliability, and realistic sample size to power a clinical trial. Importantly, WC participants’ baseline measure of peak accelerations also significantly correlated with the clinical scores of BFM right arm dystonia and disability. In this study, we wish to highlight that three of the four clinical scores showed trends of greater improvement after PSC vs. Sham than PMC vs. Sham (Supplementary Table 2, Difference column). The lack of statistical differences in clinical scores may reflect lower sensitivity of these categorical scores to detect motor changes highlighting the importance of using kinematic measures in addition to rating scales in a clinical trial.

Since the cortical gyri for PSC and PMC lie adjacent to the central sulcus, the differential stimulation response to these regions also demonstrated the cortical selectivity of our TMS effect. It is interesting that prior (9/18) studies reported a behavioral benefit after PMC-TMS (3). The majority of these studies, however, delivered 1 Hz rTMS to PMC. Therefore, the finding that PSC is a more effective stimulation site than PMC may vary by stimulation frequency. Specifically, this study showed that PSC may be more effective than PMC using 10 Hz rTMS frequency.

Future studies are needed to test if PMC or PSC may be more effective using 1 Hz rTMS frequency. Additionally, the brain state during TMS delivery may also affect the stimulation site efficacy. In this study, TMS was interleaved with writing task to prime the motor network during brain stimulation while in prior studies, TMS was interleaved with periods of rest. Since the brain state during stimulation delivery can change the plasticity inducing mechanism (long term potentiation vs. long term depression) (23), the stimulation at rest in prior studies may have induced a plasticity mechanism that may be different than the present study. Overall, our study expands the range of effective TMS parameters for adult focal hand dystonia and raises the possibility that efficacy of stimulation site may be a function of the stimulation frequency and brain state during TMS delivery.

Our second principal finding was that 10 Hz rTMS to PSC induced significant changes in subcortical connectivity in the motor network. This was an important study question to guide future refinements in therapeutic applications of TMS in dystonia, where subcortical regions such as basal ganglia and cerebellum play key roles. It is unknown whether active TMS improves behavior by weakening or strengthening brain connections. Our study showed that both weakening and strengthening of connections are present resulting in TMS-induced re-organization of the motor network. This is consistent with a prior study that applied a single session of 1 Hz rTMS to PMC and showed modification of cortical and subcortical brain regions on PET scan of primary focal dystonia participants (24). The strong association between stimulation site (PSC BOLD activity) and writing dysfluency behavior (Fig. 8) further support the potential mechanism that TMS to PSC reduces writing dysfluency behavior. Overall, the present study adds important insights on the TMS induced brain mechanism that contribute to motor behavior benefit in dystonia.

Another study finding was that the behavioral benefit after PSC-TMS was associated with strengthening of intra-cortical connectivity between somatosensory cortex and superior parietal cortex. This is consistent with a prior TMS study which showed that 1 Hz rTMS to the primary somatosensory cortex also activated the superior parietal cortex in WC dystonia (25). The superior parietal cortex is critically important for somatosensory discrimination by providing a mental model of the extremity function (26). Strengthening of the somatosensory to parietal connectivity may, therefore, be a key mechanism for developing a more accurate mental model of the hand-arm function, which in turn may improve fine motor control and reduce dysfluent writing behavior. Impairment of connectivity between the parietal cortex and somatosensory cortex were previously described in resting-state fMRIs of WC dystonia, and in multimodal imaging analyses of isolated task-specific focal dystonias (WC, musician’s dystonia, and spasmodic dysphonia) (27, 28). Impaired activation of superior parietal cortex has also been reported in cervical dystonia (29, 30). Furthermore, a prior WC study delivered a single session of cTBS to the inferior parietal lobule and showed that connectivity between the inferior parietal lobule and premotor cortex was normalized in WC compared to healthy brains (31). Collectively, prior observational and interventional studies support our study finding that the connectivity between the parietal and sensorimotor cortex may be a key target for clinical therapy in WC dystonia. Future studies should, therefore, compare the efficacy of TMS to parietal cortex, premotor cortex and primary somatosensory cortex to determine if the activation of the parietal cortex may be a more effective target for improving somatosensory dysfunction in dystonias.

To our knowledge, this is the first interventional study to identify relationships between brain connectivity and dystonic behavior in the sham-TMS condition to inform the pathophysiology. A prior study using, a systematic review of lesion induced dystonia across 359 published cases, identified that focal upper limb dystonia was most commonly caused by lesions in the basal ganglia and thalamus (32). The present study also supports these findings by demonstrating that three of the four brain-behavior connections that differentiate the effective stimulation site of PSC from noneffective stimulation sites of PMC and sham conditions are connections to or within the basal ganglia regions (caudate-substantia nigra, putamen-pallidum, putamen-substantia nigra). More importantly, the present study demonstrates that the brain connectivity pattern of subregions of the motor network are responsive to change in a direction that improves behavior after PSC-TMS compared to sham-TMS. With further validation, this brain connectivity to behavior patterns might be developed into a screening tool for future interventional trials in dystonia.

An important mechanistic question raised by these findings are whether the functional connectivity changes induced by 10 Hz PSC-TMS in the WC cohort represent a “correction” toward healthy brain connectivity pattern or a deviation from the connectivity observed in healthy individuals. While this study was not designed to assess the effects of PSC-TMS in healthy participants, a future study comparing the connectivity changes induced by 10 Hz PSC-TMS in healthy and WC subjects could differentiate between connectivity changes specific to WC pathophysiology from those that are general effects of PSC-TMS.

The brain-behavior relationships identified in this study also provide greater insight into the cerebellum’s role in dystonia. Prior neuroimaging studies observed that greater disease duration in WC participants correlated with negative cortico-cerebellar connectivity (15). Our study also identified cortico-cerebellar circuitry as potentially clinically relevant. WC participants after sham-TMS showed direct correlation between cortico-cerebellar connectivity and writing dysfluency, a relationship that was absent after PSC-TMS. Our results further support a causal role of this circuitry because TMS to PSC leads to less writing dysfluency and a significant loss of correlation between cortico-cerebellar connectivity and writing dysfluency. Our study also proposes key associations between subcortical-cerebellum and intra-cerebellar connectivity and behavior of writing dysfluency that warrant further investigation in future studies.

A key role of the cerebellum has also been described in individuals with cervical dystonia. A brain network derived from lesion network mapping and applied in rest-fMRI from cervical dystonia and healthy controls demonstrated that positive connectivity to the cerebellum and negative connectivity to the somatosensory cortex were specific markers for cervical dystonia (33). Furthermore, two weeks of cTBS-TMS to bilateral cerebellum resulted in clinical improvement in cervical dystonia (34). In DYT1-TOR1A dystonia, tractography examining cerebellar outflow tracts showed that lower measures of white matter were associated with poorer performance in a sequence learning task (35). To the extent that other dystonias may share circuit mechanisms involving reduced cerebellar connectivity, our results advance the potential for 10 Hz rTMS to PSC to be used as a corrective approach. Intriguingly, our PSC-TMS protocol may also have utility in broader neuropsychiatric conditions.

There are several limitations of the present study to discuss. First, this study consisted of a small study cohort. Nonetheless, our approach here is that small, focused studies that deeply phenotype brain-behavior relationships using a within-subject repeated study design as performed in the present study can provide key insights into brain-behavior relationships in individuals with a disease to guide clinical care (36). Second, this study only examined acute responses. Future studies are needed to examine the longevity of this TMS effect and explore whether increasing stimulation sessions prolongs the duration of behavioral effects. Third, we cannot rule out the possibility of some co-activation of adjacent cortical areas such as primary motor cortex (M1) after TMS to PMC or PSC. However, irrespective of this possibility, there was still a main effect by stimulation site on the TMS-induced behavioral and brain effects suggesting that the TMS effect at each cortical site is distinct. Fourth, a potential confounding effect of the present study is that two active stimulations and one sham condition were randomly assigned to each participant across three sequential study visits. Future studies dedicated to a single stimulation site would remove this potential confound and allow us to evaluate the longevity of each TMS condition. A fifth limitation of the present study is that we constrained analyses to the motor network because of its relevance to dystonia and to reduce multiple hypothesis testing in a study with limited group sizes. There may be significant effects in regions outside of the motor circuitry that warrant consideration if we use larger dystonia study cohorts.

In summary, identifying the optimal stimulation site for engaging and improving the abnormal motor circuit mechanisms in dystonia is a major goal necessary for effectively applying TMS-related interventions for dystonia. This study used a within-participant, sham-controlled study design in writer’s cramp dystonia, coupled with functional neuroimaging and behavior to address these unknowns. Demonstrating that TMS to PSC provides a significant behavioral benefit is a critical first step in moving TMS toward clinical therapy for dystonia. Furthermore, delineating the TMS induced corrective changes in the motor network associated with behavioral improvement in dystonia generates mechanistic hypotheses to guide future therapeutic interventions.

The pattern of brain-behavior changes observed after PSC-TMS in this study may also serve as a brain signature for a clinical response to use as a screening tool with other interventional modalities.

## Supporting information

Supplementary item

## Data availability

All data and codes are available upon reasonable request.

## Personal financial interests or professional relationships related to the subject matter but not directly to this manuscript for the preceding 3 years

NBP serves as a member of the DMRF Medical and Scientific Advisory Council. AVP is an inventor on patents and patent applications on transcranial magnetic stimulation technology and has received patent royalties and consulting fees from Rogue Research; equity options, scientific advisory board membership, and consulting fees from Ampa Health; equity options and consulting fees from Magnetic Tides; consulting fees from Soterix Medical; equipment loans from MagVenture; and research funding from Motif. SWD has received consulting fees from Neuronetics. PJM, MWL, RG, ZH, MD, ZBS, SG, ML, TKT, EW, LB, CP, JTV, LGA, HRA, AMM and NC report no relevant financial disclosures.

## Authors’ Roles

NBP, MWL, HRA, LGA, SWD, AVP and NC conceptualized the study. NBP, TKT, EW, LB collected study data. NBP, PJM, MWL, RG, ZH, MD, ZBS, SG, ML, MF and AMM performed data analysis. PT and BS performed clinician rating scales. CP and JV provided software codes to assist with data collection and analysis. MWL, and HRA critiqued the statistical analysis. MWL, SWD, AMM, AVP and NC critiqued the data analysis. NBP and NC wrote the manuscript. All co-authors reviewed and edited the manuscript.

