## Supplementary item for "Motor network reorganization associated with rTMS-induced writing improvement in writer’s cramp dystonia"

**Supplementary Methods:**

**Double blinding:** To blind study participants, participants were told that they will receive TMS at three visits, each at a different brain region and/or TMS intensity. To blind the research staff delivering TMS, the TMS scalp targeting file was void of any identifiers about the cortical location. Prior to each TMS session, the research staff stepped outside the room (and the participant was asked to close their eyes) while the study PI reoriented the AP TMS coil to placebo or active mode. The study PI then covered the TMS machine with a white sheet for the whole research visit to maintain the blinding for the research staff and study participant.

**fMRI data preprocessing:** fMRI images were preprocessed using fMRIPrep (3). fMRIPrep is an automated pipeline that performs brain extraction, head motion estimation, distortion correction, slice timing correction, intra-participant registration, and spatial normalization to the Montreal Neurological Institute (MNI) stereotaxic space (3). fMRIPrep also estimates scan quality for each fMRI run by reporting multiple regressors, including white matter, CSF, framewise displacement, and motion outliers (3).

**Prospective electric field modeling:** To perform E-field modeling, each participant's structural T1-weighted scan was utilized to construct a computational head model using the *mri2mesh* pipeline of SimNIBS (version 2.0.1 / 3.2.6) (4). The fMRI-based TMS target was then processed in the targeting and analysis pipeline (TAP) to be registered to the space of the head model. In a circular scalp area (2.5 cm radius) above the TMS target, a search over candidate coil setups was conducted in TAP to find the optimal coil setup that maximizes the directional E-field in the cortical target of interest perpendicular to the closest gyral wall. This search iterated over many candidate coil setups in steps of 1 mm and 1° increments for coil center and orientation, respectively (5-7). TAP computed an optimal coil placement for a total of 16 different hair thicknesses, prospectively ranging from 0 to 7.5 mm in intervals of 0.5 mm (7). The hair thickness of the participant at the TMS scalp target was measured on the day of the TMS visit. Out of the 16 hair thicknesses, the numerically closest to the measured value was chosen, and its associated optimal coil setup was utilized in the session with Brainsight. The coil holder was instructed to precisely maintain the optimal scalp placement of the coil relative to the TMS target by adjusting for (head) movements during the entire duration of the TMS intervention.

**Motor cortex localization and motor threshold calculation:** To identify the optimal motor cortex position of the TMS coil to activate the right first dorsal interosseous muscle, single TMS pulses were delivered while the participant was at rest. The motor cortex position was identified as the position that elicited the largest motor evoked potentials with the least intensity of the maximum stimulator output (8). The selected motor cortex position was used to calculate the resting motor threshold (RMT). Resting motor threshold (rMT) was defined as the TMS pulse intensity producing, on average an MEP of 50 μV peak-to-peak amplitude, using a maximum likelihood estimator (TMS Motor Threshold Assessment Tool, MTAT 2.0,

<http://www.clinicalresearcher.org/software.htm>)

**TMS adverse events survey:** Participants were asked to rate any adverse effects of TMS before and immediately after each TMS session. Specifically, they were asked if they experienced headache, neck pain, scalp pain, seizure, hearing impairment, cognitive impairment, trouble concentrating, mood change, or any other symptoms. Participants rated the adverse effect on an ordinal scale of absent, mild, moderate, or severe.

**TMS-induced changes in clinician-rated and participant-reported scales:** Participants were video recorded from the neck down (focused on their right arm) during the pre-TMS and post-TMS writing assessments. Movement Disorder clinicians (B.S. and P.T.) rated the videos using the Burke-Fahn-Marsden (BFM) dystonia rating scale (9) and Writer’s Cramp Rating Scale (WCRS) (10). Clinicians were blinded to the TMS conditions of the WC subjects and provided literature on the rating scales to establish concordant ratings. The two clinicians' inter-rater reliability score was measured using the intraclass correlation coefficient and observed to be 0.45 for the BFM scale and 0.76 for WCRS. WC participants also self-reported their disabilities using two rating scales: the BFM disability scale and the Arms Dystonia and Disability Scale (ADDS) (11). ADDS was scored on a 0-100% scale, with 100% denoted as normal and 0% for severe disability. Since the present study is on focal hand dystonia, for the BFM dystonia scale, clinicians were asked to rate the dystonia severity only in the right arm, and participants rated BFM disability only for handwriting. BFM handwriting disability was scored between 0-4, with 4 being severe. To measure the effect of TMS condition, the absolute change (Post-TMS minus Pre-TMS) in rating scale within participant for each TMS condition was calculated and used for statistical analyses.

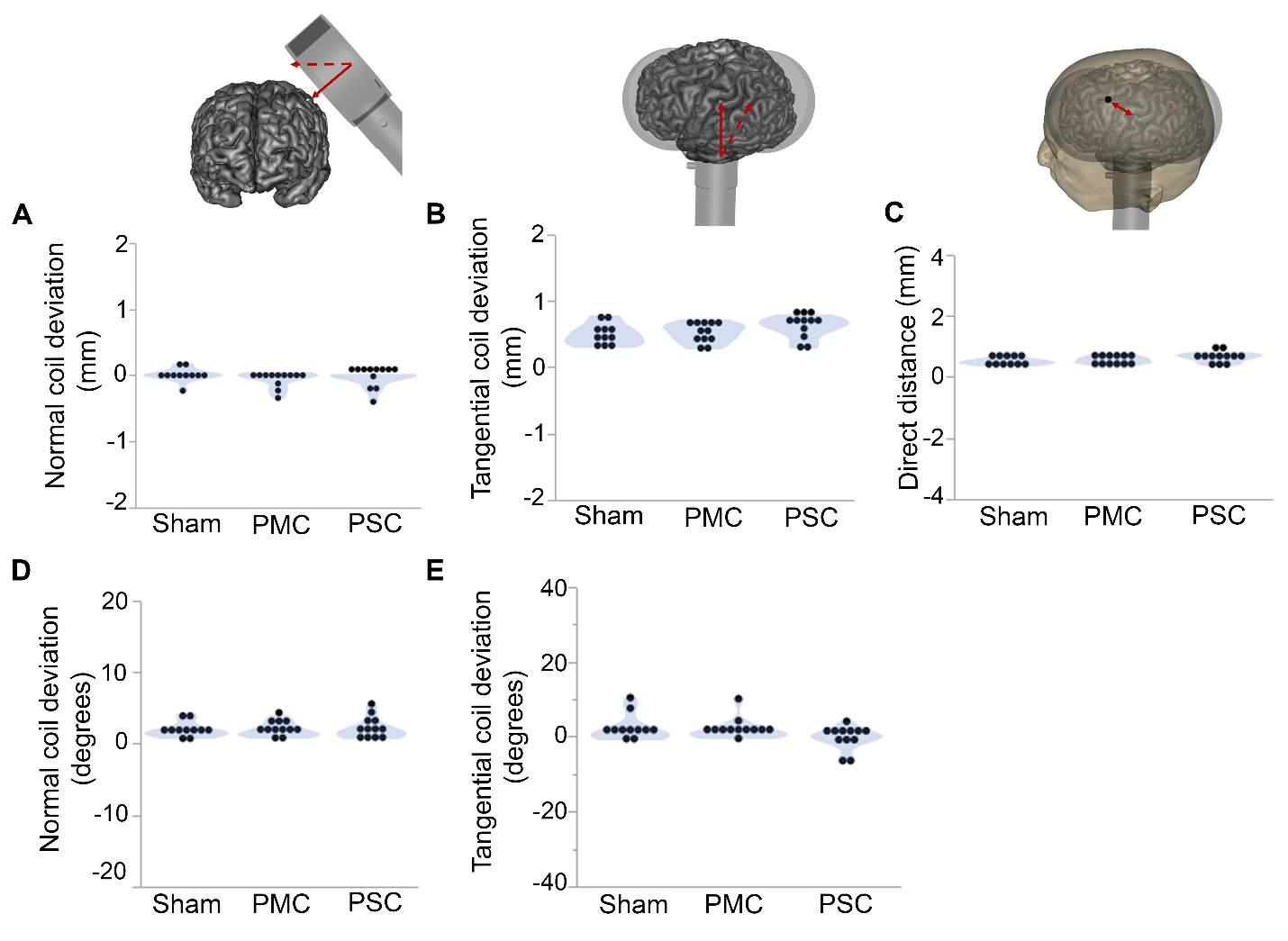

**Supplementary Figure 1:** **Minimal deviation between intended and actual TMS scalp targets.** Each graph shows the deviation in TMS coil setup defined as the difference between the intended and actual TMS coil parameters for the three TMS conditions. The deviation in TMS coil position is measured in the normal plane (A) and tangential plane (B). The direct distance between the intended TMS coil position and actual TMS coil position are also measured (C). The deviation in TMS coil orientation is also presented in the normal plane (D) and tangential plane (E). Each data point on a graph represents the mean TMS coil deviation across all four TMS blocks for each subject. Data are presented for Sham-rTMS (n = 11), 10 Hz rTMS to PMC (n = 12), and 10 Hz rTMS to PSC (n = 12). No significant differences in TMS coil deviation across the three TMS conditions were observed (MEMRM, p-value < 0.05 considered significant).

**Supplementary Table 1: A comparison of the deviations in coil position and orientation**

**for the three TMS conditions.**

| **Mean Coil Deviations** | **TMS condition** | **Difference**  **(mm or deg)** | **SE**  **(mm or deg)** | **t-ratio** | **p-value** |
| --- | --- | --- | --- | --- | --- |
| **Normal Deviation (mm)** | PSC vs. Sham | -0.07mm | 0.04 mm | -1.79 | 0.19 |
|  | PMC vs. Sham | -0.07mm | 0.04 mm | -1.63 | 0.25 |
|  | PSC vs. PMC | 0.006 mm | 0.04 mm | 0.15 | 0.98 |
| **Tangential Deviation (mm)** | PSC vs. Sham | 0.09 mm | 0.04 mm | 2.02 | 0.13 |
|  | PMC vs. Sham | 0.01 mm | 0.04 mm | 0.32 | 0.94 |
|  | PSC vs. PMC | -0.08 mm | 0.04 mm | -1.74 | 0.21 |
| **Direct Distance (mm)** | PSC vs. Sham | 0.10 mm | 0.05 mm | 1.96 | 0.14 |
|  | PMC vs. Sham | 0.02 mm | 0.05 mm | 0.4 | 0.91 |
|  | PSC vs. PMC | -0.08 mm | 0.05 mm | -1.60 | 0.26 |
| **Normal Deviation (deg)** | PSC vs. Sham | 0.12 deg | 0.24 deg | 0.53 | 0.85 |
|  | PMC vs. Sham | 0.02 deg | 0.24 deg | 0.09 | 0.99 |
|  | PSC vs. PMC | -0.1 deg | 0.23 deg | -0.44 | 0.89 |
| **Tangential Deviation (deg)** | PSC vs. Sham | -2.73 deg | 1.42 deg | -1.91 | 0.15 |
|  | PMC vs. Sham | -0.38 deg | 1.42 deg | -0.27 | 0.95 |
|  | PSC vs. PMC | 2.34 deg | 1.39 deg | 1.67 | 0.23 |

**SE:** Standard Error

**Supplementary Table 2: Comparison of TMS-induced changes in clinician and subject rating scales.**

| **Rater** | **Rating Scale** | **TMS condition** | **Difference**  **(#)** | **SE**  **(#)** | **t-ratio** | **p-value** |
| --- | --- | --- | --- | --- | --- | --- |
| **Clinicians** | **BFM dystonia**  **right hand** | PSC vs. Sham | 0.07 | 0.13 | 0.54 | 0.59 |
|  |  | PMC vs. Sham | 0.05 | 0.13 | 0.42 | 0.68 |
|  |  | PSC vs. PMC | -0.02 | 0.13 | -0.13 | 0.90 |
|  | **WCRS movement score** | PSC vs. Sham | 1.24 | 0.67 | 1.85 | 0.08 |
|  |  | PMC vs. Sham | 0.03 | 0.67 | 0.05 | 0.96 |
|  |  | PSC vs. PMC | -1.2 | 0.66 | -1.83 | 0.08 |
| **WC subjects** | **BFM writing score** | PSC vs. Sham | 0.03 | 0.19 | 0.18 | 0.86 |
|  |  | PMC vs. Sham | -0.05 | 0.19 | -0.27 | 0.79 |
|  |  | PSC vs. PMC | -0.08 | 0.18 | -0.46 | 0.65 |
|  | **ADDS total score** | PSC vs. Sham | 2.28 | 2.22 | 1.03 | 0.31 |
|  |  | PMC vs. Sham | 2.74 | 2.22 | 1.24 | 0.23 |
|  |  | PSC vs. PMC | 0.46 | 2.14 | 0.21 | 0.83 |

**SE:** Standard Error

**Reference Citations**
